# The Modular Actigraphy Platform: A Data Science Solution for Processing High-Resolution Time Series Sensor Data for Sleep and Physical Activity Assessment

**DOI:** 10.1101/2025.11.19.25340572

**Authors:** Pin-Wei Chen, Dipriya A. Pillai, Michael S. Campagna, Catherine M. Avitabile, Sara King-Dowling, Stephanie L. Mayne, Scott M. Haag, Jonathan A. Mitchell

## Abstract

**Introduction:** Wearables with proprietary scoring protocols are typically used to assess sleep and physical activity, but the field is shifting to wearables with raw sensor data accessible and open-source scoring methods to enhance rigor and reproducibility. The data infrastructure to process raw sensor data in clinical research is underdeveloped; we therefore developed the Modular Actigraphy Platform (MAP).

**Methods:** MAP is a cloud-based computational platform that processes high-resolution time series sensor data to derive sleep and physical activity metrics. It was engineered to be modular, providing flexibility in data processing and enabling the seamless integration of open-source sleep and physical activity scoring methodologies as they become available (currently, GGIR and MIMS processing algorithms have been integrated). A structured Software Development Life Cycle (SDLC) approach was used to guide the development of MAP, with a multi-level testing framework consisting of unit testing (verification of modules), integration testing (interaction among modules), and system testing (validating specifications). Following these foundational tests, we then completed user acceptance testing in two phases - alpha (17 test files) and beta (686 files from 4 pediatric cohorts) to assess processing performance.

**Results:** For beta testing, MAP leveraged up to 60 CPU cores and 500 GiB of memory. The pre-processing module was the most computationally demanding and was more efficient in MAP compared to offline processing (up to 8 CPU cores and 23.2 GiB of memory). For example, the preprocessing GGIR part 1 container was completed at a speed of 0.29-0.49 minutes per file (1.6-2.9 times faster than offline processing) and the pre-processing MIMS container was completed at a speed of 0.49-4.66 minutes per file (2.4 to 14.0 times faster than offline processing).

**Conclusion:** MAP is an efficient computational platform that integrates open-source scoring algorithms to efficiently process raw sensor data for wearable sleep and physical activity estimation in clinical research and is available through the Children’s Hospital of Philadelphia’s Research Institute.

## Introduction

Sleep and physical activity have a profound impact on health in childhood. Population-based studies have shown that insufficient sleep^1^ and physical inactivity^2^ in childhood are highly prevalent and are associated with multiple risk factors - such as obesity^3-5^ and hypertension^6-8^ - that are precursors to cardiovascular disease (CVD), cancer, Alzheimer’s disease, and other leading causes of morbidity and early mortality^9^. Further, sleep and physical activity are increasingly being investigated in the context of improving outcomes for children with medical conditions, such as autism^10^, cancer^11^, chronic kidney disease^12^, congenital heart disease^13,14^, hypertension^6^, migraine^15^, neurodevelopmental disorders^16^, and diabetes^17,18^. However, assessment of these behaviors is limited by the lack of standardized methods to derive estimates with rigor and reproducibility.

Wearable devices are commonly used to assess sleep and physical activity in children. This passive approach captures extensive biometric time series data which can be processed to provide sensor-based estimates of sleep and physical activity in the community setting over multiple days, weeks or months. However, most wearable device protocols leverage motion sensor data that is stored in a proprietary data format. This approach has at least one of the following limitations. 1) The estimates are device dependent. 2) Single-user license software installed on a local computer creates a data silo that limits collaborations and integration with other data repositories. 3) Proprietary software can be updated with limited ability for version control. 4) Proprietary devices and software can be discontinued rendering protocols obsolete. 5) The need for highly trained personnel is costly and limits scalability. 6) Storing data on the device requires manual upload, increases risk of data loss, and is restrictive for remote patient monitoring.

Recognizing these limitations, the field is moving to use wearable devices that collect and provide raw sensor data, ideally with Bluetooth and Wi-Fi facilitated remote data capture, and open-source scoring methods. However, the data infrastructure and computational workflows needed to manage and process the high-resolution time series sensor data are underdeveloped. Indeed, most open-source algorithms are accessed and utilized through GitHub repositories, and this requires advanced data science expertise and fluency in multiple programming languages, with extensive time and effort needed to resolve defects in the source code depending on how well it is maintained. Also, most open-source algorithms provide single-step solutions (e.g., sleep/wake estimation only) and lack the steps needed for complete end-to-end data processing (i.e., raw data pre-processing -- non-wear detection -- sleep/wake estimation -- sleep period identification -- and physical activity estimation).

To help enhance the data infrastructure and computational workflows for high-resolution time series data processing, we have developed the Modular Actigraphy Platform (MAP). This platform integrates open-source algorithms that have utility in raw acceleration data processing for sleep and physical activity estimation. Version 1 included the GGIR package^19^ to provide complete end-to-end processing for sleep and physical activity assessment, and version 2 added the Monitor Independent Movement Summary (MIMS) pre-processing algorithm to enhance the physical activity scoring options^20^. The platform is designed to be accessible to a broad range of clinical research teams, with or without advanced data science expertise. The overarching goal of MAP is to provide a user-friendly computational tool to democratize access to the most innovative wearable device methodologies for sleep and physical activity assessment using raw sensor data. In this paper, we describe how MAP was designed and developed and demonstrate the performance of this tool.

## Methods

### Overview of MAP

MAP is a cloud-based computational platform that processes high-resolution time series sensor data to derive sleep and physical activity metrics (Figure 1). This platform processes raw tri-axial acceleration data from multiple wearable devices. MAP was engineered to be modular, providing flexibility in data processing and enabling the seamless integration of open-source sleep and physical activity scoring methodologies as they become available (currently, GGIR^19^ and MIMS^20^ processing algorithms have been integrated). MAP was also designed to be cloud-agnostic, capable of running on major cloud platforms like Google Cloud Platform (GCP; Alphabet, Mountain View, CA), Amazon Web Service (AWS; Seattle, WA), or Microsoft Azure (Redmond, WA). For our deployment, we leveraged GCP. A team of investigators with expertise in pediatric sleep and physical activity research collaborated with data scientists, computer scientists, and platform engineers to design, build, secure and maintain MAP. Definitions of key components of MAP are given in Table 1.

**Table 1.**
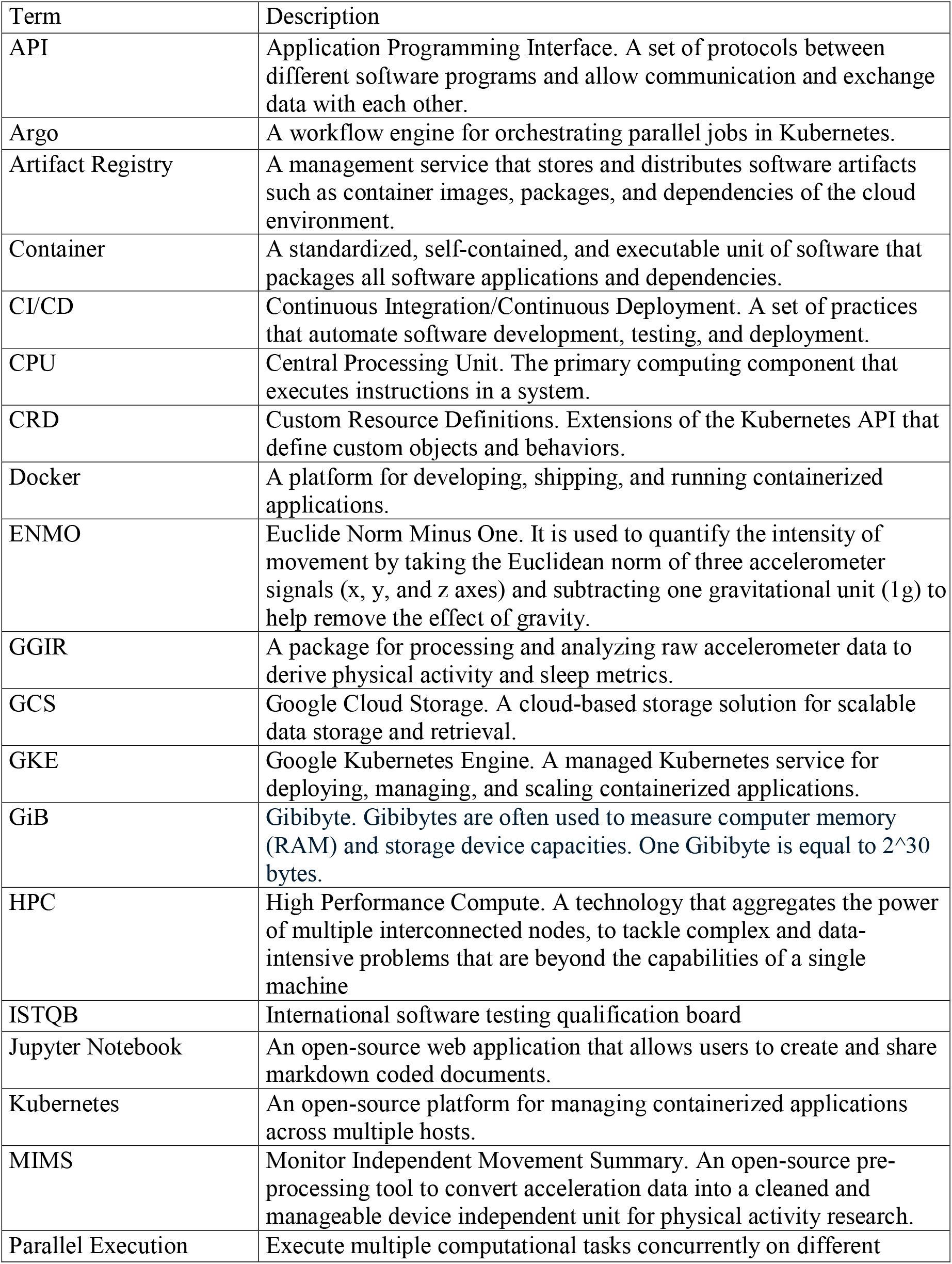
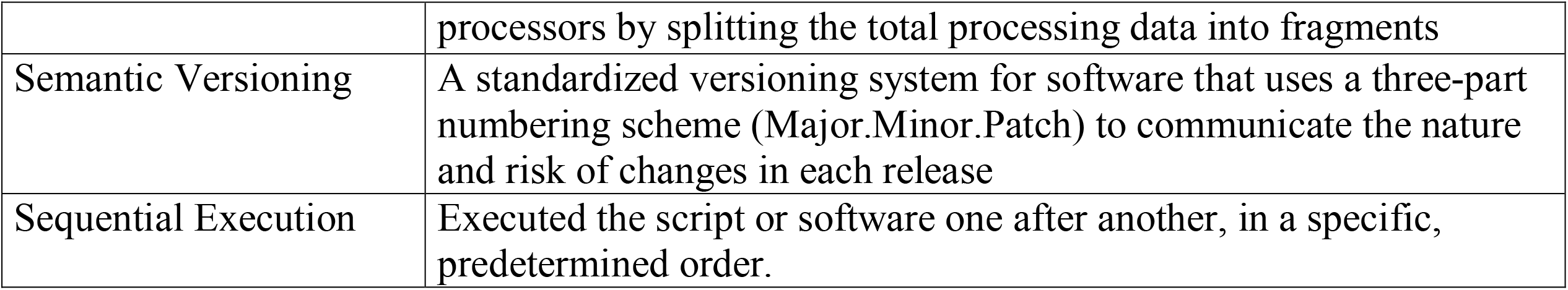
Glossary of terms related to key features of MAP.

**Figure 1.**
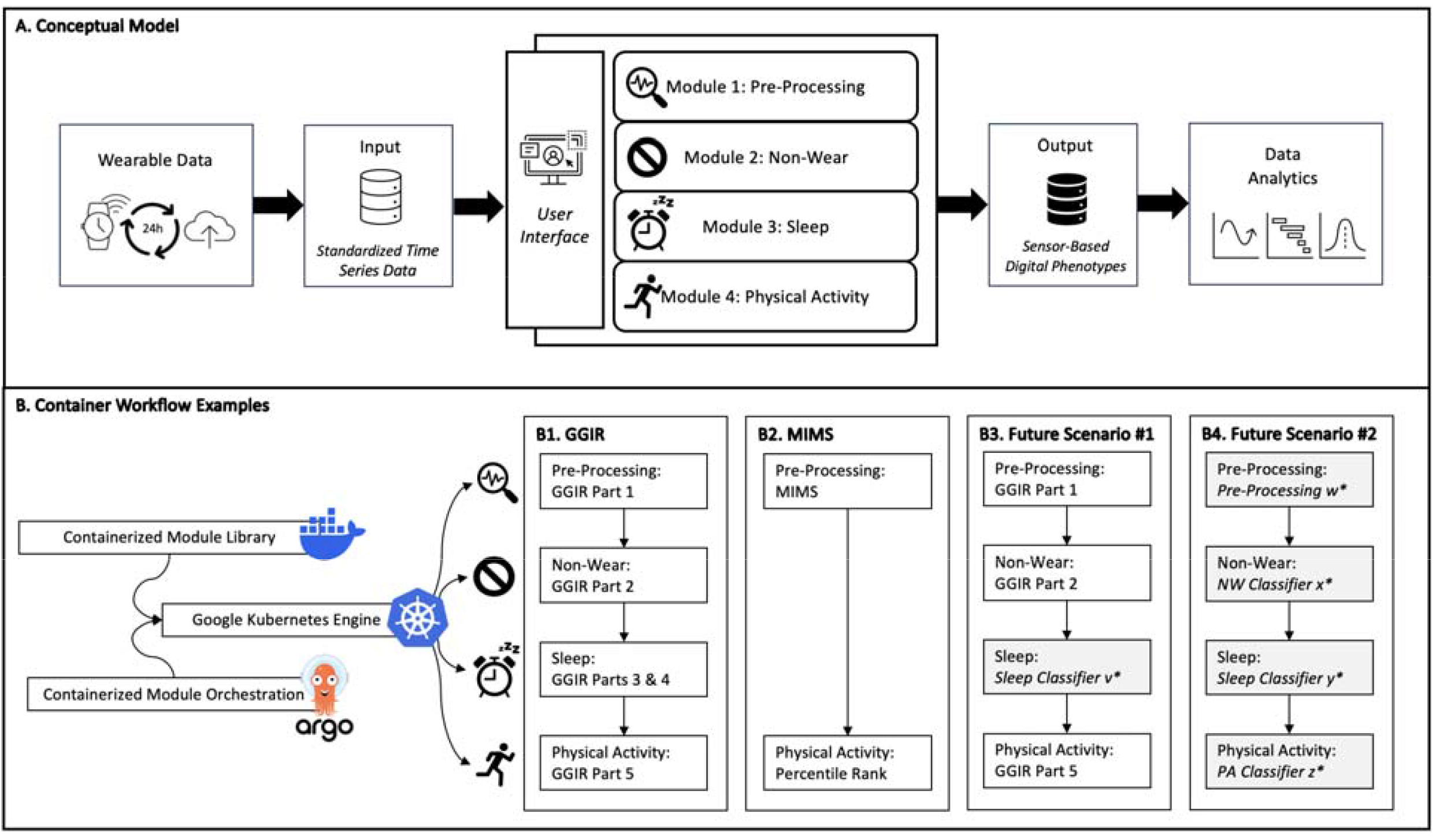
Visual illustration of the Modular Actigraphy Platform (MAP): conceptual model (panel A) and technical workflow examples (panel B). Panel B1: Standard GGIR end-to-end processing. Panel B2: Standard MIMS processing. Panel B3: Near-term future scenario leveraging GGIR but with substituting in a sleep classifier developed using statistical learning or machine learning methods. Panel B4: Long-term future scenario envisioning a workflow using novel pre-processing, novel non-wear detection algorithms, novel sleep detection algorithms, and novel physical activity scoring algorithms. Containerized modules are stored as Docker images in an algorithm library. Argo orchestrates containerized module execution and manages workflow sequences. Google Kubernetes Engine (GKE) runs containerized modules independently in a scalable compute environment.

### Wearable Sensor Data Input

MAP processes tri-axial accelerometer data collected at high-resolution sampling rates (e.g., 50Hz). We have processed raw acceleration data acquired from wrist-worn ActiGraph (Ametris, Pensacola, Florida) and GENEActiv (Activinsights, Kimbolton, UK) devices. The raw tri-axial accelerometer data can be processed using device specific compressed file formats (i.e.,.gt3x and.bin) using GGIR. However, these file formats do not operate for the MIMS pre-processing methods; we therefore used the packages *GGIRread*^21^ and *read*.*gt3x*^22^ to create a custom script to convert the device specific file formats to a unified.csv format with standardized timestamps and column headers to allow for seamless data processing initiation.

### Modular Design

MAP is implemented as a series of containerized modules, with each module encapsulated as a Docker image. This architecture contrasts with traditional sequential step-based designs that are typical when processing sensor data, where all processing stages are tightly coupled within a single processing flow. Containerizing each module allows alternative algorithms to be applied at any processing stage simply by substituting, or running in parallel, the alternative container. This modular design offers several key benefits:

- *Independent Updates and Replacements*: Any module can be modified, upgraded, or replaced through adaptation at the container level without requiring changes to the rest of the platform.
- *Algorithm Flexibility:* Researchers can select or test different processing or scoring algorithms at a given modular stage by replacing the container.
- *Environment Isolation:* Each container includes its own dependencies, preventing version conflicts and ensuring reproducibility across the platform.

MAP’s architecture supports both sequential and parallel execution of modules, enabling efficient resource utilization and scalability. By decoupling processing stages through containerization, MAP provides a flexible, extensible, and reproducible platform for wearable sensor data processing. A user-interface allows for project specific configurations to be specified through an interactive Jupyter notebook^23^, which includes step-by-step instructions for setup and execution. The first two versions of MAP integrated the GGIR^19^ and MIMS^20^ processing tools and this interface allows users to select one or both approaches. When GGIR is selected, the settings can be customized directly in the notebook to accommodate study-specific requirements. To illustrate the functionality of this modular design, current and future processing workflows are provided in Figure 1 (Panel B).

### Security

Security and stability are maintained through comprehensive vulnerability scans and proactive software maintenance practices. Google’s Vulnerability Scanner^24^ is used to scan every built image pushed to the Artifact Registry, identifying vulnerabilities categorized as critical, high, medium, or low. The MAP engineering team promptly addresses all critical and high vulnerabilities before deployment. Additionally, this team ensures that all software components remain up to date with the latest versions, minimizing exposure to known threats and maintaining compatibility with new updates and features. To further enhance security, regular image scanning and the use of minimal base images help reduce the attack surface. Containers are deployed using Google Kubernetes Engine (GKE)^25,26^ with built-in runtime protection, and critical patches are applied as soon as they become available to maintain a secure and stable environment.

### Version Control

MAP employs version control procedures to ensure consistent source code management, facilitate collaboration among contributors, and maintain transparency throughout software development. GitHub serves as the primary platform for version control, enabling seamless tracking of code changes, issue management, and peer review through pull requests. MAP adheres to the principles of Semantic Versioning^27^, using the MAJOR.MINOR.PATCH convention to distinguish the scope of changes. Major versions indicate significant updates that introduce incompatible changes, minor versions add new features in a backward-compatible manner, and patch versions apply backward-compatible bug fixes and minor improvements. This systematic versioning approach allows users to understand the nature of changes introduced in each release, ensuring informed decision-making regarding updates and deployments.

GitHub Actions are integrated into the MAP repository to automate the build. Continuous integration and deployment (CI/CD) pipelines are configured to run security scans and only verified builds are pushed to the Google Artifact Registry, ensuring that deployed containers meet quality and security standards. Overall, this approach follows the industry development standard and ensures traceability, reliability, and transparency.

### MAP Version 1: GGIR and Module Containers

GGIR is a widely used R package for processing raw acceleration data from wearables to derive estimates of sleep and physical activity. Developed and maintained by the data science company *Accelting*, GGIR is openly available through the Comprehensive R Archive Network (CRAN) as an integrated, step-based package. Its technical details are well documented in prior publications and online resources^19^, and its performance for estimating sleep and physical activity - including in pediatric populations - has been assessed in multiple studies^28-37^. We integrated GGIR into the first version of MAP because it provides complete, end-to-end functionality for converting raw acceleration data into standardized sleep and physical activity metrics.

For MAP, we restructured the GGIR computational workflow into four distinct containerized modules. In the pre-processing module, GGIR’s part 1 methods are applied to remove artifacts from high-resolution acceleration data (i.e., excessively high and low frequencies) and to adjust for background gravity. The non-wear detection module incorporates the GGIR part 2 algorithm to identify extended periods of minimal or no acceleration, estimating non-wear time. The sleep scoring module applies GGIR parts 3 and 4 to classify epochs of sleep and wake (part 3) and to apply a heuristic guider to detect primary sleep periods (part 4). This module generates key sleep health metrics such as sleep duration, onset and offset timing, wake after sleep onset, sleep efficiency, and the sleep regularity index. The physical activity scoring module integrates GGIR part 5 to estimate time spent in sedentary behavior and across different physical activity intensities, along with the intensity gradient algorithm to capture overall physical activity levels.

### MAP Version 2: MIMS Container

The Monitor Independent Movement Summary (MIMS) algorithm was developed by John et al.^20^ and converts raw tri-axial acceleration data into an acceleration summary called MIMS-units. This is a pre-processing method analogous to GGIR part 1. The first step of this algorithm processes raw acceleration data to harmonize the sampling rate (“interpolation”); the second step corrects instances in the time series data where the dynamic range has been exceeded (extrapolation); third, acceleration artifacts that are not representative of human movement are filtered out, to minimize the contribution of vibrations and tremors, and to remove the gravity vector (“bandpass filtering); and fourth, the single axis and triaxial data are aggregated using an area under the curve approach (as opposed to Euclidian norm) to capture total movement and movement intensity. The aggregated epoch length can be set to a desired specificity (e.g., 5, 30 or 60 seconds)^20^. We had to make a minor fix to the MIMS algorithm script available from the open-source GitHub page to correct a coding error, and we modified to add in batch data processing and parallel processing capability in MAP.

The MIMS container was added to MAP because there are normative reference data to convert MIMS units into age and sex percentiles to quantify the total volume of physical activity. The reference data were developed using acceleration data from the National Health and Nutrition Examination Survey (NHANES), providing a normative ranking standard using a representative sample of the national U.S. population^38^.

### MAP Development and Testing Procedures

A structured Software Development Life Cycle (SDLC) approach was used to guide the development of MAP. This cycle includes the following steps: requirement gathering, design, implementation, testing, deployment, and maintenance. This was used to ensure that MAP evolved in a systematic manner, with iterative improvements over successive versions and clear traceability from research needs to deployment^39-41^.

Complementing the SDLC, we established a multi-level testing framework consisting of black box unit testing (verification of individual modules), integration testing (ensuring interaction among modules), system testing (validating the complete pipeline against specifications), and user acceptance testing (validation with end users)^42^. This aligns with internationally recognized standards^41,43^.

Unit, integration, and system testing were conducted first to verify the correct operation of the individual MAP containerized modules and their interoperability. This confirmed that the containerized modules functioned independently and in combination. In addition, dependencies and interactions were evaluated to ensure proper integration with external components, including Google Cloud Storage (GCS) buckets for data management, Google Artifact Registry for container images, and the Google Container Analysis/Vulnerability Scanner for image security alongside MAP’s internal modules.

Following this foundational verification, we then completed user acceptance testing. This was performed across two phases (alpha and beta). The goal of alpha testing was to evaluate MAP at the platform level through end-to-end sample runs in a development environment. The goal of beta testing was to further evaluate functionality, performance, scalability, and robustness in the production environment using real-world datasets. In the results section, we present the processing performance of the containerized modules of MAP across alpha and beta testing phases. For comparison, offline data processing using a virtual machine and the same acceleration files are included.

## Results

### Alpha Testing

This testing stage was done in a development environment, with limited resources, using 17 test acceleration data files that were generated by research staff at CHOP, totaling 4.9 GB with data spanning from 1 to 14 days (Table 2).

**Table 2.** Acceleration data files used for alpha testing.

| Test Phase | Dataset | File Format | No. Files | Total Size (MB) | Average (MB) | SD (MB) |
| --- | --- | --- | --- | --- | --- | --- |
| Alpha | Run 1 | .bin | 8 | 3,112.5 | 389.1 | 320.3 |
| Alpha | Run 1 | .gt3x | 6 | 566.1 | 94.4 | 71.2 |
| Alpha | Run 1 | .csv | 3 | 1,348.9 | 449.6 | 57.1 |
| Alpha | Run 2 | .bin | 50 | 18,432.0 | 348.6 | 292.2 |
| Alpha | Run 2 | .gt3x | 50 | 4,915.2 | 96.8 | 66.9 |

The first run was executed with up to 10 CPUs and 100 GiB RAM allocated. In version 1, the end-to-end completion of the GGIR processing took 11.4 minutes, with 63% of time dedicated to the pre-processing (GGIR part 1 container: 7.2 minutes, Table 3). In version 2, the pre-processing MIMS container – using the same data and resources – was completed in 23.9 minutes (Table 3). The MIMS pre-processing step was therefore 3.3 times slower than the GGIR part 1 pre-processing step.

**Table 3.** Alpha testing performance metrics of MAP.

| Module | Container | Run | No. Files | Total Size (GB) | CPU (Cores) | Memory (GiB) | Time (min) | Speed (min/file) |
| --- | --- | --- | --- | --- | --- | --- | --- | --- |
| Preprocessing | GGIR Part 1 | 1 | 17 | 4.9 | 10 | 100 | 7.2 | 0.42 |
|  |  | 2 | 100 | 25 | 12 | 120 | 45.3 | 0.45 |
| Non-wear | GGIR Part 2 | 1 | 17 | 4.9 | 10 | 100 | 1.3 | 0.07 |
|  |  | 2 | 100 | 25 | 12 | 100 | 7.6 | 0.08 |
| Sleep | GGIR Part 3 & 4 | 1 | 17 | 4.9 | 10 | 100 | 1.6 | 0.09 |
|  |  | 2 | 100 | 25 | 12 | 100 | 10.8 | 0.11 |
| Physical Activity | GGIR Part 5 | 1 | 17 | 4.9 | 8 | 32 | 1.3 | 0.07 |
|  |  | 2 | 100 | 25 | 12 | 100 | 8.5 | 0.08 |
| Preprocessing | MIMS | 1 | 17 | 4.9 | 10 | 100 | 23.9 | 1.40 |
|  |  | 2 | 100 | 25 | 12 | 100 | 127.0 | 1.27 |

In a second run, to stress test, the 17 files were expanded by random duplication to 100 files totaling 25 GB. Allocating 10 CPUs and 100 GiB RAM, the processing failed due to insufficient memory. We therefore increased to allocate 12 CPU cores and 120 GiB RAM. Using these resources, the end-to-end completion of the GGIR processing in version 1 took 72.2 minutes (Table 3); again, 63% of the time was dedicated to the pre-processing step (GGIR part 1 container: 45.3 minutes, Table 3). In version 2, the second stress test run for the pre-processing MIMS container was completed in 127.0 minutes (Table 3). The MIMS pre-processing step was therefore 2.8 times slower than the GGIR part 1 pre-processing step.

Overall, development environment testing proved that versions 1 and 2 of MAP were end-to-end operational. This level of testing also provided initial quantification of resources needed to overcome insufficient memory.

### Beta Testing

For this stage, we leveraged real-world acceleration data files collected in the Sleep and Growth Study 2 (S-Grow2), the Teen Neighborhood Activity Patterns and Sleep (Teen NAPS) study, the Physical Activity and Cancer Experience (PACE) studies, and the Pulmonary Hypertension (PH) study. Full cohort details are given in the Supplemental File.

In brief, participants in S-Grow2, Teen NAPS, and PH were asked to wear a 3-axis GENEActiv accelerometer on their non-dominant wrist for 14 days, with the devices configured to collect acceleration data collected at a sampling rate of 30Hz (PHP) or 50Hz (S-Grow2 and Teen NAPS). Participants in the PACE studies wore an ActiGraph GT9X accelerometer on their non-dominant wrist for 14 days, with the raw acceleration data collected at a sampling rate of 30Hz. Collectively, these four studies provided 686 raw acceleration files (.bin and.gt3x) equaling 437.7 GB (Table 4).

**Table 4.** Beta Testing Performance Metrics.

| Study Details | Module | Container | MAP w/ HPC |  |  |  | Offline Computing |  |  |  | MAP : Offline Speed |
| --- | --- | --- | --- | --- | --- | --- | --- | --- | --- | --- | --- |
|  |  |  | CPU (Cores) | Memory (GiB) | Time (Min) | Speed (Min/file) | CPU (Cores) | Memory (GiB) | Time (Min) | Speed (Min/file) |  |
| S-Grow2<br>N=323 .bin files<br>243.3 (SD: 0.06) GB | Pre-Processing | GGIR Part 1 | 60 | 120 | 128.9 | 0.40 | 8 | 23.2 | 210.0 | 0.65 | 1.63 |
|  | Non-Wear | GGIR Part 2 | 40 | 100 | 56.5 | 0.17 | 8 | 23.2 | 11.5 | 0.04 | 0.20 |
|  | Sleep | GGIR Parts 3/4 | 40 | 100 | 66.8 | 0.21 | 8 | 23.2 | 72.0 | 0.22 | 1.08 |
|  | Physical Activity | GGIR Part 5 | 40 | 100 | 56.4 | 0.17 | 8 | 23.2 | 36.9 | 0.11 | 0.65 |
|  | Pre-Processing | MIMS | 20 | 500 | 444.0 | 1.37 | 8 | 23.2 | 6,007.1* | 18.60* | 13.53 |
| Teen NAPS<br>N=176 .bin files<br>135.0 (SD: 0.09) GB | Pre-Processing | GGIR Part 1 | 60 | 120 | 87.0 | 0.49 | 8 | 17.2 | 215.7 | 1.23 | 2.48 |
|  | Non-Wear | GGIR Part 2 | 40 | 100 | 29.5 | 0.17 | 8 | 17.2 | 2.3 | 0.01 | 0.08 |
|  | Sleep | GGIR Parts 3/4 | 40 | 100 | 37.3 | 0.21 | 8 | 17.2 | 49.5 | 0.28 | 1.33 |
|  | Physical Activity | GGIR Part 5 | 40 | 100 | 35.2 | 0.20 | 8 | 17.2 | 48.6 | 0.28 | 1.38 |
|  | Pre-Processing | MIMS | 20 | 500 | 156.0 | 0.89 | 8 | 23.2 | 2,182.3 | 12.40 | 13.99 |
| PACE<br>N=127 .gt3x files<br>12.9 (SD: 0.04) GB | Pre-Processing | GGIR Part 1 | 60 | 330 | 45.4 | 0.36 | 8 | 23.2 | 133.0 | 1.05 | 2.93 |
|  | Non-Wear | GGIR Part 2 | 40 | 100 | 7.6 | 0.06 | 8 | 23.2 | 6.8 | 0.05 | 0.89 |
|  | Sleep | GGIR Parts 3/4 | 40 | 100 | 12.6 | 0.10 | 8 | 23.2 | 54.4 | 0.43 | 4.32 |
|  | Physical Activity | GGIR Part 5 | 40 | 100 | 9.1 | 0.07 | 8 | 23.2 | 16.4 | 0.13 | 1.80 |
|  | Pre-Processing | MIMS | 20 | 500 | 592 | 4.66 | 8 | 23.2 | 1,411.0 | 11.11 | 2.38 |
| PH<br>N=60 .bin files<br>42.5 (SD: 0.05) GB | Pre-Processing | GGIR Part 1 | 60 | 120 | 17.5 | 0.29 | 8 | 23.2 | 40.1 | 0.67 | 2.29 |
|  | Non-Wear | GGIR Part 2 | 40 | 100 | 7.5 | 0.13 | 8 | 23.2 | 2.6 | 0.04 | 0.35 |
|  | Sleep | GGIR Parts 3/4 | 40 | 100 | 7.9 | 0.13 | 8 | 23.2 | 12.9 | 0.22 | 1.63 |
|  | Physical Activity | GGIR Part 5 | 40 | 100 | 7.4 | 0.12 | 8 | 23.2 | 7.3 | 0.12 | 0.99 |
|  | Pre-Processing | MIMS | 20 | 500 | 133.0 | 2.22 | 8 | 23.2 | 776.7 | 12.94 | 5.84 |

\*Due to the amount of time and resources needed to run the offline MIMS analysis, we sampled 5 files from the 323 files and calculate the average of the time needed per file. Then we estimate how much time will that need in 323 files.

A production environment, with more resources than the development environment, was used to process data files separately for each study. Initially, we allocated up to 12 CPUs and 120 GiB of memory for each project and increased resources as needed to avoid insufficient memory errors. For GGIR processing, most studies required up to 60 CPUs and 120 GiB memory for part 1 pre-processing, and up to 40 CPUs and 100 GiB memory for parts 2-5 (Table 4). The overall GGIR processing times for S-Grow2, Teen NAPS, PACE, and PHP using MAP were 309 minutes, 189 minutes, 75 minutes, and 40 minutes. The GGIR part 1 pre-processing container was the most computationally demanding for all four studies, taking 41-61% of the total processing time (Table 4). In contrast, the overall offline processing times using up to 8 CPU cores and 23.2 GiB of memory were slower for all four studies (Table 4). For example, the GGIR part pre-processing container was 1.6 to 2.9 times slower compared to MAP (Table 4).

For MAP version 2 beta testing of the pre-processing MIMS container, up to 20 CPUs and 500 GiB memory were allocated to provide sufficient resources (Table 4). The processing times for S-Grow2, Teen NAPS, PACE, and PHP were 444 minutes, 156 minutes, 592 minutes, and 133 minutes (Table 4). This was 2.4 to 14.0 times faster than offline processing (Table 4). Compared to GGIR part 1 preprocessing, MIMS preprocessing was 3.4 to 13.0 times slower using MAP (although direct comparison is not possible, due to allocated compute differences; Table 4).

Batch processing was needed to overcome memory failures when using the MIMS pre-processing container in MAP (e.g., the S-Grow2 files were processed in batches of 100, 100 and 123 files). For offline processing, more extensive batching and splitting of files was required to successfully process the data using the MIMS algorithm (one file was processed at a time, and each file was split into one day chunks).

## Discussion

MAP is a secure, efficient, and adaptable computational tool for processing raw wearable sensor data to generate estimates of sleep and physical activity in clinical research. Leveraging a cloud-based computing architecture, MAP integrates open-source scoring algorithms and orchestrates data processing workflows for sensor data, providing a scalable method for sleep and physical activity estimation from wearables with rigor and reproducibility. With a focus on the need for assessing sleep and physical activity in childhood, the methods integrated have been shown to have acceptable validation for use in pediatric research (as well as adult research). The key innovation of MAP is the modular design, with independent containerized modules. We used a software development lifecycle approach and a multi-level testing framework to design and test each version of MAP. We demonstrated the operational performance using test data files and real-world pediatric data files, with MAP processing being more computationally efficient compared to offline processing. Overall, MAP democratizes access to advanced data science methods to process raw sensor data for sleep and physical activity assessment.

GGIR was selected as the first algorithm to be integrated as containerized modules into MAP because it has all the steps needed for complete end-to-end raw acceleration data processing for both sleep and physical activity assessment^19^. The singular step-based MIMS algorithm followed to add to the pre-processing functionality of MAP and provide an additional physical activity scoring option^20,38^. There are several additional step-based open-source algorithms that exist and are being considered for module-specific integration into MAP. We are also leading and partnering with investigators to develop novel sleep and physical activity scoring methods and MAP is an ideal platform through which such methods can be tested and implemented. MAP is therefore an adaptable tool that will continue to expand its functionality over time.

The current version of MAP helps to overcome many of the known limitations of wearable device scoring protocols for sleep and physical activity estimation. 1) MAP integrates open-source algorithms and processes raw tri-axial acceleration data from different types of devices, therefore generating estimates that are device independent. 2) MAP is cloud-based capable of integrating with other data repositories (e.g., Clinical Data Warehouses) and so is not a data silo or single-user licensed software that operates on a local computer. 3) With semantic version control and security measures in place, MAP defaults to the latest software versions but can also draw upon archived versions as needed. 4) Proprietary devices and software can be discontinued rendering protocols obsolete; MAP addresses this limitation by providing a user-friendly platform to process raw sensor data that is not contingent upon proprietary data file formats. 5) Accessing and applying most open-source algorithms requires data science expertise; MAP makes the application of such algorithms more accessible to clinical research teams with or without a dedicated data scientist. 6) The current version of MAP in production does not have automated data upload capabilities. However, developmental work is ongoing to add this capability and will allow MAP to be used in studies requiring timely and efficient data processing at scale (e.g., a processing solution for raw wearable sensor data collected daily for months/years).

The first versions of MAP are accessible through secure, project-specific computational labs supported by the Children’s Hospital of Philadelphia (CHOP) Research Institute’s Arcus Program. These labs have MAP enabled, with storage for the raw acceleration data provided, and Arcus supported platform engineer oversight ensuring that MAP is operating per the security measures described. The added benefit is that Arcus labs can securely integrate with CHOP’s electronic health record data via our Clinical Data Warehouse and other institutional databases (e.g., the Arcus Geographic Information Systems Geocoded Database), therefore, facilitating innovative wearable clinical informatic research focused on sleep and/or physical activity. For investigators outside of CHOP, MAP is accessible within an Arcus supported computational lab through collaboration with a CHOP investigator, and their processed data can be exported for further analysis within their own research program. We are actively working to enhance the portability of MAP so investigators can more readily access this tool without the need for such collaboration. This is feasible but will require resources to implement and maintain since the modular workflow requires platform engineering oversight.

MAP is well suited for processing raw acceleration data files collected from a range of study designs. A typical observational research study, using a cross-sectional or longitudinal design, will ask participants to provide 1-2 weeks of wearable device data. Per our beta testing, the pre-processing GGIR part 1 and MIMS containers were the most computationally demanding when processing data from such study designs, with the MIMS container being especially computationally demanding. Indeed, we found that processing raw data files with 7 or more days of data using the MIMS algorithm was challenging using a local computing resource environment and required loading data in batches for processing. With 8 CPUs and 23.6 GiB memory of computing power with parallel processing, we found that it took an average of 14 minutes (SD=3) to process a single file with 14-days of data collected at 30-50Hz sampling rate. This aligns with the original paper, where John et al. reported an approximate time of 18 min to process one approximately 7-days file (80 Hz sampling) on a Dell Precision 7810 workstation (64 GB memory, 24-core 3.0 GHz CPU) without parallel processing^20^. Using MAP with up to 20 CPUs and parallel processing, the MIMS container was completed within 2 minutes (SD=1.67) on average. This is approximately 6 times faster and suggests that MAP is capable of processing raw data at larger scales, over multiple months. Indeed, we intend to use MAP to process raw acceleration data collected in a pediatric sleep and exercise intervention trials with up to 365 days of data per participant being led at CHOP^44^.

For our deployment of MAP, we used GCP. Cloud resource assignments fluctuate (e.g., CPU type, resource contention, or network latency), so MAP processing speed will be variable between runs. Unlike an on-premises system with a fixed CPU, cloud jobs can land on different processor generations or experience variable resource sharing, which affects timing. For example, the ultramem-961 GiB node (≈40 vCPUs, ∼2.2–2.6 GHz base, ∼961 GiB RAM) prioritizes large memory capacity, while the newer c3-highcpu-88 node (≈88 vCPUs, 3.0 GHz sustained all-core turbo, ∼176 GiB RAM) is optimized for compute performance. Consequently, MAP stages that are CPU-bound (e.g., GGIR preprocessing and MIMS analysis) tend to run faster on the C3 high-CPU nodes despite their smaller memory footprint and more RAM would not equate to a faster tun. Overall, even with expected cloud-side variability, MAP processing remained faster than the offline implementation.

There are limitations of MAP. First, MAP integrates open-source software, and our intent is to use the source code as provided by the developers. This has proven to be the case for GGIR, which is well maintained and updated regularly. However, we modified the original MIMS source code to fix a one-line error and to accommodate the need for batch processing. Note, this modification did not alter the key processing steps and final data output. Second, new releases to GGIR occur at a relatively high frequency, sometimes with errors that are later fixed. Our built-in security requires the use of the latest software releases, and if errors are present, this impacts the operation of MAP. If this occurs, MAP reverts to the most recent stable version we have verified through alpha and beta testing. Third, we have focused on the processing of raw acceleration data; however, in future versions of MAP we will expand to include additional raw sensor data, such as light and photoplethysmography (PPG) sensor data to enhance sleep and physical activity scoring options.

In conclusion, sleep and physical activity are key behaviors that need to be measured with rigor and reproducibility using wearable devices using secure, adaptable, and scalable methods. MAP contributes to this goal by providing a cloud-based computational tool to aid raw sensor data processing without extensive data science expertise.

## Data Availability

All data produced in the present work are contained in the manuscript.

## Acknowledgments

MAP was developed at the Children’s Hospital of Philadelphia’s Research Institute. The Clinical and Translational Science Award at CHOP (CTSA@CHOP) oversees the development and dissemination of MAP through the Digital Health Innovation Core. The CHOP Arcus team provides computer science and platform engineering expertise to host, operate and securely maintain MAP; whereas the Data Science and Biostatistics Unit (DSBU) at CHOP provides data science expertise to help maintain and update the functionality of MAP and the biostatistics expertise to analyze the phenotypic data generated. Finally, a CHOP-based scientific steering committee of faculty and staff with expertise in sleep and physical activity research provides guidance on the scientific direction of MAP. The development of MAP was supported by grants from the National Institutes of Health. The CTSA@CHOP is supported by NIH/NCATS UL1 TR001878. The Sleep and Growth Study 2 (S-Grow2) is supported by NIH/NICHD R01 HD100421. The Pulmonary Hypertension studies are supported by NIH/NHLBI K23HL150337. The Teen NAPS project is supported by NIH/NHLBI K01 HL155860. The PACE studies are supported by grants from the University of Pennsylvania Institute for Translational Medicine and Therapeutics (NIH/NCATS UL1 RR024134), the CHOP Center for Pediatric Clinical Effectiveness and the American Institute of Cancer Research. Arcus is supported by the CHOP Research Institute.

## Supplemental File: Beta Testing Cohort Descriptions

The Sleep and Growth Study 2 (S-Grow2) is an observational study of typically developing adolescents with the primary goal of determining of sleep impacts bone health. Participants were enrolled when they were in in 7^th^ grade (12-13y) and are followed-up at 1-year intervals in 8^th^ and 9^th^ grades. Data collection is ongoing, so only the baseline data are included in the cross-sectional beta testing analysis. All data were collected during a school term between 2021-2024 in the southeastern PA and southern NJ region. Participants wore a 3-axis GENEActiv accelerometer on their non-dominant wrist for 14 days, with the raw acceleration data collected at a sampling rate of 50Hz. In the current performance test, we only include the visit 1 data. The total file size was 243.27 GB (Table 4). The study protocol was approved by the Institutional Review Board at the Children’s Hospital of Philadelphia (IRB: 20-17328).

The Teen Neighborhood Activity Patterns and Sleep (Teen NAPS) Study is a cross-sectional observational study of typically developing adolescents with the goal of determining how daily home and neighborhood environmental exposures impact sleep outcomes. Participants were aged 15-18 and lived and attended school in Philadelphia, PA. All data were collected between 2022 and 2024. Participants wore a 3-axis GENEActiv accelerometer on their non-dominant wrist for 14 days, with the raw acceleration data collected at a sampling rate of 50Hz. The total file size was 134.14 GB (Table 4). The study protocol was approved by the Institutional Review Board at the Children’s Hospital of Philadelphia (IRB: 21-018763).

The Physical Activity and Cancer Experience (PACE) studies included data collected across 3 projects aimed at determining real-time predictors and benefits of physical activity behavior among adolescents and young adults impacted by cancer (both on and off treatment). Participants were between the ages of 14-24 and enrolled from CHOP oncology clinics between 2022-2024. Participants wore a ActiGraph GT9X accelerometer on their non-dominant wrist for 14 days, with the raw acceleration data collected at a sampling rate of 30Hz. The data recording for gt3x is setup in a way that the recording can be longer than 14 days. The total file size was 13.25 GB (Table 4). Because the recordings of this study were set to record data until download, many of the monitors can be recording up to 30 days. The study protocols were approved by the Institutional Review Board at the Children’s Hospital of Philadelphia (IRB: 17-014087, 22-019674, and 23-020794).

The pulmonary hypertension (PH) pilot study consists of thirty youth with pediatric pulmonary hypertension and thirty healthy control patients, enrolled in a cross-sectional observational study of physical activity, musculoskeletal health, and functional status. Participants between the ages of 8 and 18 years were enrolled from the CHOP Pulmonary Hypertension Program and outpatient general pediatric programs between 2021 and 2024. Participants wore a GENEActiv accelerometer on their non-dominant wrist for 14 days, with the raw acceleration data collected at a sampling rate of 30 Hz. The total file size was 43.48 GB (Table 4). The study protocol was approved by the Institutional Review Board at the Children’s Hospital of Philadelphia (IRB: 20-017827).

